# Concerns, Priorities, and Pathways: Healthcare Provider Perspectives on Implementing Tuberculosis Preventive Treatment for People with HIV in Indonesia and the Philippines

**DOI:** 10.64898/2026.07.13.26357999

**Authors:** Prashant Kulkarni, Laura Steiner, Alieen Gianan-Gascon, Candice Eula Lamigo, Bianca Joyce Sornillo, Ruth Anne Hechanova-Cruz, Anna Maureen Dungca-Lorilla, Aljira Fitya Hapsari, Retnosari Hardaningsih, Mira Yulianti, Anshari Saifuddin Hasibuan, Evy Yunihastuti, Priyanka Raichur, Rossana Ditangco, Jonathan E Golub, Christopher J Hoffmann, Mark Donald C. Reñosa

**Affiliations:** Johns Hopkins Centre for Infectious Diseases in India (CIDI), Pune, India; Johns Hopkins University, School of Medicine, Baltimore, USA; Research Institute for Tropical Medicine – Department of Health, Muntinlupa City, Philippines; Universitas Indonesia, Jakarta, Indonesia; CiptoMangunkusumo General Hospital, Jakarta, Indonesia; Heidelberg Institute of Global Health, Faculty of Medicine and University Hospital, Heidelberg University, Heidelberg, Germany

**Keywords:** Tuberculosis, HIV, People living with HIV, TPT implementation, Healthcare providers

## Abstract

**Background:** Tuberculosis preventive therapy (TPT) is a cornerstone intervention for reducing TB incidence among people living with HIV, yet its integration into routine care remains inconsistent in high-burden settings. While policy frameworks are well established, less is known about how frontline healthcare workers (HCWs) operationalize TPT within constraint health systems. Thisstudy explores healthcare workers’ (HCWs) perspectives on TPT implementation in Indonesia and the Philippines using normalization process theory (NPT), to understand how TPT is (or fails to be) embedded in everyday clinical practice.

**Methods:** Weconducted ten focus group discussions and four in-depth interviews with HCWs across major TB/HIV treatment centers in Manila, Philippines and Jakarta, Indonesiabetween June to December 2023. Data were analyzed using reflexive thematic analysis, followed by interpretative mappingontofour NPT constructs: coherence, cognitive participation, collective action and reflexive monitoring.

**Results:** TPT implementation was characterized by a persistent gap between policy intent and routine practice. Under *coherence*, HCWsdescribed fragmented knowledge and limited exposure to standardized protocols, undermining confidence in TPT delivery. *Cognitive participation* was constrained by a doctor-centric model and high staff turnover, weakening shared ownership of TPT. In *collective action*, operational fragilities, including weak documentation systems, supply interruptions, and poor inter-level coordination, resulted in inconsistent delivery. Through *reflexive monitoring*, HCWs identified patient concerns around side effects and pill burden, alongside provider concerns regarding diagnostic uncertainty and fears of missing active TB. Across settings, HCWs actively proposed system-oriented solutions, including SOP standardization, team-based training, digital reminders, and strengthened care continuity mechanisms.

**Conclusions:** TPT implementation is not limited by awareness alone but by the absence of routinized systems that support consistent practice. Strengthening implementation requires shifting from individual-dependent delivery toward system-enabled integration, through standardized protocols, distributed workforce engagement, embedded monitoring mechanisms, and uninterrupted drug supply systems. Interventions that align with everyday clinical workflows and reinforce collective responsibility across cadres are critical to closing policy and practice gap.

## BACKGROUND

Tuberculosis (TB) remains a major global health challenge and continues to be a leading cause of death among people living with HIV (PLHIV), accounting for an estimated 161,000 deaths in 2023 alone(1). PLHIV remain disproportionately vulnerable to TB-related morbidity and mortality worldwide (2,3).For these high-risk populations, TB preventive therapy (TPT) is a critical intervention that reduces TB incidence, prevents progression to active disease, and lowers TB related mortality.The World Health Organization’s (WHO) consolidated guidelines recommend that PLHIV who are unlikely to have active TB should receive TPT as part of a comprehensive HIV care package(4,5).Despite robust evidence and global endorsement, the integration of TPT in routine HIV careremains uneven,particularlyin high-burden, resource-constrained settings(6–8).Despite the availability of national and international guidelines, integrating TPT into routine HIV care remains challenging, as successful implementation depends on how healthcare workers (HCWs) interpret, operationalize, and sustain these recommendations within existing service delivery systems(6,7,9,10).

Implementation gaps are not solely the result of resource limitations but reflect deeper misalignments between policy design and service delivery realities. Health system constraints includingfragmented workflows, unclear task distribution, inconsistent drug supply, and weak integration of TPT into routine HIV care, interact with provider-level uncertainties and patient concerns to undermine consistent uptake(4,6,8). Health system constraints including fragmented workflows, unclear task distribution, inconsistent drug supply, and weak integration of TPT into routine HIV care interact with provider-level uncertainties and patient concerns to undermine consistent uptake.In this context, TPT implementation is no longer a question of whether it is recommended, but rather how it can be effectively operationalized in routine clinical practice(9,11,12).

These implementation gaps are often negotiated through the day-to-day work of frontline HCWs, who must interpret, adapt, and operationalize TPT within existing healthcare systems. To better understand these lived realities, we drew on Normalization Process Theory (NPT), a sociological framework that examines how new interventions become embedded into routine practice(13). NPT focuses onfour interrelated constructs: coherence (how HCWs make sense of the intervention), cognitive participation (how stakeholders engage and commit), collective action (how practices are enacted), and reflexive monitoring (how interventions are evaluated and adapted)(13,14). This theoretical framing is particularly valuable in capturing the dynamic, everyday work of HCWs as they negotiate the integration of TPT into existing systems(13).By applying this lens, we move beyond asking *what*hinders or enables TPT,andtoward understanding *how* TPT becomes(or fails to become)a part of everyday clinical practice.

While prior studies have identified barriers related to system readiness, guideline dissemination, and patient concerns, there remains a paucity of contextual research exploring HCWs’ perspectives on TPT implementation, particularly in Southeast Asia(10,15,16).In Indonesia and the Philippines, two countries grappling with high TB/HIV co-infection rates and ongoing health system reforms, little is known about the priorities, adaptations, and strategies that frontline HCWs employ to deliver TPT effectively. While our earlier work has identified structural and behavioral barriers to TPT uptake(17), the present study advances this understanding by unpacking the dynamic processes through which healthcare workers interpret, operationalize, and adapt TPT implementation within real-world health systems.By centering HCWs’ experiences, this study offers actionable insights into the concerns,priorities and pathways shaping the integration of TPT into routine HIV care.

## MATERIALS AND METHODS

### Study Design and Setting

We conducted aqualitative study to explore the HCWs’ perspectives with TPT implementation across two high-burden TB/HIV settings in Southeast Asia. Data collection was carried out at the Research Institute for Tropical Medicine (RITM) in Muntinlupa city, Philippines, and CiptoMangunkusumo General Hospital (Cipto) in Jakarta, Indonesia, both critical HIV treatment hubs in their respective countries. RITM, the Philippines’ largest HIV center, manages approximately6,000 new HIV cases annually, whereasCiptoservesas Indonesia’s national referral hospital for HIV, treating approximately500 new cases yearly. These settings provide ideal contexts for examining TPT implementation challenges and facilitators within established healthcare systems serving high-riskpopulations.

### Data Collection and Sampling Strategy

HCWs were purposively selected to ensure representation across key cadres, includingdoctors, nurses, counselors (only for the RITM study site), andpharmacists. Participants were identified in collaboration with site leads based on their involvement in HIV and TB care.

Between June and December 2023, ten focus group discussions (FGDs) and four in-depth interviews (IDIs) were conducted with HCWs involved in HIV and TB care. IDIs were conducted with individuals whose schedules or roles precluded participation in FGDs. The FGDs and IDIs were led by trained Filipino-speaking and Bahasa Indonesia-speaking interviewers. Data collection instruments included an information sheet, an interview guide, informed consent forms, and debriefing forms, all pilot-tested for clarity and contextual relevance. Additional probes and follow-up questions were incorporated to explore HCWs’ experiences, implementation challenges, and perceptions of patient engagement with TPT. FGDs and IDIs lasted between 45 and 120 minutes, with systematic debriefings conducted to refine lines of inquiry, facilitate reflexive discussion, and assess data saturation. On the basis of the specificity of the sample and the depth of discussions regarding TPT implementation, data saturation was achieved after ten FGDs and four IDIs.

### Data analysis

All FGDs and IDIs were audio recorded, transcribed verbatim in Filipino or Bahasa Indonesia, and then translated into English by trained transcribers following qualitative research standards(18). Transcripts were reviewed for accuracy by study teams at each site. Transcripts, interviewer notes, and debriefings were managed and coded using MAXQDA24 (VERBI Software, 2024).

Reflexive thematic analysis was used to systematically identify, analyze, and interpret themes emerging from the data. Themes were generated through an inductive analytic process grounded in the data, rather than being predefined by a theoretical framework. Once the themes were developed and refined, the NPT was introduced as a framework to better understand the social processes influencing TPT implementation. NPT constructs, such ascoherence, cognitive participation, collective action, and reflexive monitoring,provided a structured way to interpret findings but did not dictate the initial coding or theme development. This approach ensures methodological rigor by demonstrating that the NPT emerged from, rather than dictated, the data analysis. This alignment between the data-driven themes and NPT constructs facilitates a deeper understanding of how contextual factors impact the implementation process, ultimately leading to more effective strategies for enhancing TPT adoption in various settings. By emphasizing the interplay between theoretical frameworks and empirical data, we identified specific concerns and priority areas that influence TPT implementation, paving the way for tailored interventions that address unique challenges in different contexts.The data were coded on the basis of four NPT constructs (see **Table 1**).

**Table 1.** Operational definition of constructs of Normalization Process Theory (NPT) (13)

| NPT Constructs | Operational Definition |
| --- | --- |
| **Coherence** | Exploring how HCWs make sense of TPT within their roles and healthcare settings. |
| **Cognitive Participation** | Examining engagement and collaboration among stakeholders in implementing TPT. |
| **Collective action** | Identifying challenges and facilitators in operationalizing TPT practices, including resource allocation, training, and guideline adherence. |
| **Reflexive monitoring** | Investing participants' reflections on the impact and effectiveness of TPT, including strategies for improvement. |

Coding was conducted iteratively by three researchers (PR, MDCR and LS), with regular analytical discussions to explore interpretations, challenge assumptions, and refine themes.Rather than seeking consensus, our process emphasized reflexivity and analytical depth, consistent with reflexive thematic analysis.

The findings were mapped to NPT constructs to provide a structured understanding of HCWs’ experiences and to inform actionable recommendations for improving TPT implementation. This approach facilitated a comprehensive analysis of both systemic and individual-level factors influencing TPT prescription, uptake and adherence.The application of NPT as a secondary analytic lens facilitated a theoretical exploration of emerging themes while preserving the flexibility of the inductive coding process, thereby enhancing both empirical foundation and conceptual richness.

## RESULTS

### Participant characteristics

Of63 HCWs approached, 52 agreed to participate – 26 from the Philippines and 26 from Indonesia. The 11HCWs who declined participation citedscheduling conflicts (n=7) or did not specify a reason (n=4). The study participants were a mix of men, women and HCW cadres. Most had over 5 years working in the TB field(see **Table 2**).

**Table 2.** Demographic profile of HCW participants (n=52).

| **Characteristics** | **Philippines**<br>**(n=26)**<br>**(n, %)** | **Indonesia (n=26)**<br>**(n, %)** |
| --- | --- | --- |
| **Sex** |  |  |
| Male | 14, 54% | 8, 31% |
| Female | 12, 46% | 18, 69% |
| **Age group** |  |  |
| <30 years | 5, 19% | 3, 12% |
| 31-40 years | 14, 54% | 14, 54% |
| 41-50 years | 5, 19% | 6, 23% |
| >50 years | 2, 8% | 3, 12% |
| **Healthcare provider classification** |  |  |
| Medical Doctor | 8, 31% | 12, 46% |
| Nurse | 5, 19% | 9, 35% |
| Pharmacists | 2, 8% | 5, 19% |
| Peer counselor | 7, 27% | 0, 0% |
| Others: Support staff | 4, 15% | 0, 0% |
| **Number of years working in HIV/Tuberculosis field** |  |  |
| <1 year | 4, 15% | 0, 0% |
| 1-2 year/s | 3, 12% | 5, 19% |
| 3-5 years | 5, 19% | 5, 19% |
| >5 years | 14, 54% | 16, 62% |
| **Prescribes TPT\*** |  |  |
| Yes | 8, 31% | 12, 46% |
| No | 18, 69% | 14, 54% |
| **Provides counseling services** |  |  |

| Characteristics | Philippines<br>(n=26)<br>(n, %) | Indonesia (n=26)<br>(n, %) |
| --- | --- | --- |
| Yes | 24, 92% | 26, 100% |
| No | 2, 8% | 0, 0% |

Acrossboth settings, HCWs described that the implementation of TPT was characterized by a conflict between the expectations set by policies and the practical challenges encountered in operations. Although national guidelines established TPT as a standard component of HIV care, its implementation continued to rely on individualinitiative, informal practices, and creative solutions. This led to differences in the understanding, implementation, and maintenance of TPT across various facilities.

Using the Concern-Priority-Pathwayanalytic structure mappedonto NPT constructs, we identified how HCWs move from recognizing implementation challenges to proposing actionable pathways for integrating TPT into routine care (see **Table 3**). Across all four NPT constructs – coherence, cognitive participation, collective action, and reflexive monitoring - HCWsdescribed structural and adaptive processes.

**Table 3.** Summary of HCWs’Concerns, Priorities and Pathways for TPT Implementation, Mapped to NPT Constructs.

| NPT Construct | Concerns | Identified Priorities | Proposed Pathways |
| --- | --- | --- | --- |
| **COHERENCE**<br>(Making sense of TPT) | • **Limited Awareness:** HCWs had limited awareness of national TPT guidelines and institutional SOPs.<br>• **Knowledge Gaps:** HCWs lacked sufficient knowledge, particularly regarding side-effects and TPT eligibility criteria. | • **Standardization and Clarity:** standardization and clarity in TPT regimens to generate implementation coherence.<br>• **Concise, Uniform Instructions:** concise, uniform instructions on drug choices (e.g., 3HP vs 6H), dosages, and duration. | • **Strengthen Knowledge:** Consistent messaging and a need to strengthen HCWs' knowledge.<br>• **Clearer SOPs:** Clearer SOPs integrated into Electronic Medical Record (EMR) systems and regular briefings. |
| **COGNITIVE PARTICIPATION**<br>(Building engagement) | • **Reliance on Doctors:** TPT counseling and initiation relied heavily on doctors, limiting | • **Collaborative Patient Education:** Prioritizing collaborative patient education.<br>• Involving **nurses** and | • **Team-Based Orientation:** Team-based TPT orientation sessions and inter-cadre communication |
|  | engagement from other cadres.<br>• **Risk of Knowledge Loss:** Staff rotation systems (posed a risk to the sustainment and transfer of core TPT knowledge. | **pharmacists** in TPT counseling. | platforms.<br>• **Reinforcement:** Ensure multiple cadres reinforce TPT messages across various care touchpoints. |
| **COLLECTIVE ACTION (Delivering in routine practice)** | • **Inconsistent Documentation/Lack of SOP:** Absence of a formal SOP for documenting TPT provision led to inconsistency in recording.<br>• **Coordination Gaps:** Lack of reliable communication and effective interdisciplinary coordination.<br>• **Logistical Barriers:** Mention of staff shortages and stock interruptions. | • **Process Formalization:** Ensuring effective interdisciplinary coordination is critical to embed TPT into routine care.<br>• **IT Support:** Implementing IT-based reminders/formal SOP for consistent TPT status documentation. | • **Formalized Processes/IT Systems:** Implement IT-based reminders to alert doctors to a patient's TPT status.<br>• **Supply Chain Management:** Improve patient access by ensuring a six-month supply of TPT drugs.<br>• **Continuity of Care:** Strengthen linkages and communication between primary healthcare providers and hospitals.<br>• **Patient support groups:** Development of patient support group through social reinforcement |
| **REFLEXIVE MONITORING (Reflecting and adapting)** | • **Patient Reluctance/Side Effects:** reluctance to initiate TPT due to concerns about side effects (e.g., numbness, nausea) and the pill burden. | • **Enhanced Counseling:** Strengthen counseling that emphasized the benefits of TPT.<br>• **Side-Effect Mitigation:** Prescribe vitamin B-complex alongside isoniazid to reduce side-effects. | • **Enhanced Counseling:** Strengthen counseling on TPT benefits.<br>• **Adaptation:** Proactively suggesting Vitamin B-complex for side-effect management. |

### Coherence: Making Sense of TPTin Practice

A central finding was limited internalization of TPT guidelinesat the facility level. Although national policies endorsed TPT, many HCWs reported minimal exposure to institutional standard operating procedures (SOPs) and inconsistent clarity regarding regimen selection, eligibility criteria, and side-effect management. One nurse explained, “*Whatever SOP is there, in the case of TPT, we haven’t actually been exposed to it*.”(Nurse, Philippines)

Respondents reported less knowledge regarding management of side-effects and differentiation between regimens (3HP vs 6H) which further reduced their confidence in initiating TPT discussions: “*We are not informed about the complications of TPT or how to manage them, so we face challenges in addressing these issues when patients come to us*.” (Medical doctor, Indonesia)

In response, HCWs prioritized clearer and more standardized, concise, and uniform guidance across cadres. Proposed pathways included integratingSOPs into electronic medical records (EMRs), conducting regular interdisciplinary briefings, and reinforcingconsistent messaging across doctors, pharmacists, nurses, and counselors. In both study sites, informal reinforcement already occurred: “*Our staff, including the pharmacists, they reinforce… the importance of Isoniazid preventive treatment*” (Medical doctor, Indonesia). HCWs mentioned that these reinforcements emerge but have incomplete integration, dependent largely on their initiatives rather than system-level standardization.

### Cognitive Participation: Building Engagement and Shared Responsibility

Engagement in TPT implementation was constrained by a predominantly doctor-centric model, where counselling and initiation responsibilities rested largely with physicians, limiting involvement from other cadres. Frequent staff rotations, particularly among contractual nurses, further contributed to discontinuity and loss of institutional knowledge.

> *“Well, the doctor should understand who should use 3HP, 6H, those are the two kinds of drugs”* (Medical doctor, Indonesia).

HCWs identified the need to shift towards shared responsibility, prioritizing greater involvement of nurses and pharmacists in patient education and follow-up, as well as sustained engagement across cadres despite workforce instability: “*TPT must be explained again. Maybe outside, the nurse, while taking blood pressure, immediately asks which TPT candidate is eligible*” (Medical doctor, Indonesia).

Encouraging teamwork among HCWs is also reported to be essential for effectively identifying and monitoring TPT candidates. As noted, “*Yes, actually, the patient is exposed more often and remembers more often. Oh, I see. At the front, counseling has been given…*” (Health provider, Phillipines), highlighting the value of reinforcing messages across multiple care touchpoints.

To address this, they proposed team-based TPT orientation sessions and strengthening inter-cadre communication platforms to maintain continuity of knowledge. Additionally, reinforcing TPT messages at multiple care touchpoints, such as triage, consultation, and pharmacy, was seen as essential to enhance patient understanding, improve recall, and support adherence. This distributed approach was perceived as critical to reducing reliance on a single cadre and fostering collective ownership of TPT delivery.“*Again, the only usual question is when to take it, is it okay to take it with other medications, and the duration*” (Pharmacist, Indonesia), underscoring the importance of clear guidance in supporting patient compliance.

> *“Actually, it can be overcome every time after that all the doctors type and then keep writing again that this patient has received TPT. The only thing is that there is no SOP for us to write that. Second, we have a rotation system here, people change again so you cannot mean as long as it is not needed, you cannot expect that; So there is a risk that if it is a year later, the next doctor who accepts this patient will not know whether this patient has received TPT or not.”*(Medical doctor,Philippines)

### Collective Action: Operational Fragility in Routine Practice

The operationalization of TPT within routine clinical workflows revealed significant system-level fragility, characterized by the absence of formal SOPs for documentation, weak coordination across cadres and levels of care, staff shortages, and inconsistent drug supply. They described how frequent stock interruptions undermined confidence in prescribing TPT and disrupted continuity of care.These challenges resulted in fragmented implementation, including inconsistent recording of TPT provision, poor tracking of patient status, and disruptions in continuity of care.

> *“We organized outreach activities about prescribing because there are doctors who tend to mix up the medicines. He [patient] should not have been prescribed 3HP; instead, he was prescribed 3HP or INH. Well, the doctor should already understand, who should use 3HP or 6H, since these are the two kinds of drugs.”* (Pharmacist, Indonesia)
>
> *“However, we have nurses there who are in Contract of Service. Therefore, the turnaround of that is pretty fast. So they will… sometimes they will leave…”* (Medical doctor, Philippines)
>
> *“INH [Isoniazid] still doesn’t exist. We don’t know where INH is.”*(Pharmacist, Indonesia)

In response to these operational challenges, HCWs emphasized the need to formalize routine processes and strengthen coordination mechanisms to ensure consistent and reliable TPT delivery. Key priorities included establishing clear documentation protocols, improving interdisciplinary communication, ensuring uninterrupted drug supply, and strengthening linkages between primary and tertiary care to maintain continuity of care.

> *“Actually, thist can be overcome every time if all the doctors type and then keep writing again that this patient has received TPT. The only thing is that there is no SOP for us to write that”* (Medical doctor, Indonesia).
>
> *“What it means is a system to remind doctors, to provide notification that the patient has or has not received TPT”* (Medical doctor, Indonesia).

Participants proposed IT-based reminders and formalized documentation SOPs to track TPT provision.They also emphasized ensuring a six-month drug supply, strengthening coordination between primary and tertiary care and building social support networks to enhance adherence. Ensuring continuous drug supply through multi-month dispensing was also highlighted as a key strategy to reduce logistical barriers. Furthermore, strengthening referral and communication systems between primary and tertiary care levels was seen as essential for maintaining continuity, while the development of patient support groups was suggested to enhance adherence through social reinforcement. Together, these pathways aimed to embed TPT more effectively within routine systems and reduce reliance on ad hoc practices.

> *“So our pharmacy in CIDTRAM [satellite pharmacy for HIV patients] uh… they dispense medications to the patients good for 6 months already”* (Nurse, Philippines).
>
> *“The support group is good, like if cancer patients join… it creates a sense of group affinity”* (Medical doctor, Indonesia).

### Reflexive Monitoring: Reflecting and Adapting based on Experience

Reflections on implementation highlighted patient-level and system-level barriers, including reluctance to initiate TPT due to perceived side effects, pill burden, and misconceptions, as well as role ambiguity between doctors and nurses in managing TPT.

> *“Your palm and sole of foot will numb, so to counteract the side effects of isoniazid, you need to give them vitamin B complex.”* (Nurse, Philippines)

HCWs prioritized strengthening patient-centred counselling to address these concerns and improve acceptance, alongside clarifying roles within the care team.

> *“The decision to initiate IPT [Isoniazid preventive therapy] rests with the doctor, as nurses can only advise but must follow the doctor’s judgment. Once IPT is prescribed, nurses are responsible for educating patients on proper medication usetaking isoniazid on an empty stomach (30 minutes before or 2 hours after meals) for effective absorption, understanding side effects, and the importance of taking vitamin B complex alongside it. Nurses also provide discharge instructions, reinforce adherence, and monitor whether patients are consistently taking and completing the six-month IPT course.”*(Support staff, Philippines)

To address these challenges, they proposed enhancing counselling to emphasize the benefits of TPT and provide clear guidance on managing side effects, including the routine use of vitamin B-complex alongside isoniazid. Clear delineation of responsibilities, where doctors lead initiation and nurses support counselling, adherence monitoring, and follow-up,was also emphasized. In addition, HCWs recommended integrating TPT indicators into hospital data systems and linking TPT performance to accreditation and evaluation metrics to strengthen institutional accountability. These strategies were seen as critical for enabling ongoing learning, improving patient adherence, and ensuring sustained implementation.

> *“Add to accreditation assessment points. Therefore, for example, if a hospital has an application, right? Usually, when it’s time for an evaluation, all the registry systems will be fixed straight away”* (Medical doctor,Philippines).

## DISCUSSION

Ourstudy offers an in-depth account of how HCWs in Indonesia and the Philippines experience, negotiate, and sustain TPT implementation within routine HIV care. UsingNPT as an interpretativelens, we found that TPT implementation is not merely shaped by the presence or absence of policy, but by everyday work required to make it understandable, actionable, and sustainable in practice. Unlike prior studies that primarilyquantify TPT uptake or catalogue structural and behavioral barriers, our findings frame implementation as a dynamic, relational, and contextually embeddedprocess in which frontline HCWs actively interpret policy, respond to operational constraints, and develop practical strategies to sustain delivery(6,10,19). Our findings also demonstrate how HCWs move beyondrecognizing implementation challenges to identifyingwhat matters most and proposing feasible, system-oriented solutions. This framinghighlights implementation as an active and iterative process of translation, through which frontline providers convertsystemic constraints into concrete actions across the continuum of care.

Although TPT has been recommended within the National TB Program for PLHIV since 2014 in Indonesia and 2020 in the Philippines, implementation remains suboptimal in both countries (20,21). Our findings suggest that this gap is not simply a matter of insufficient policy guidance, but of limited routinization: TPT has not been fully embedded into the knowledge systems, division of labor, workflows, and evaluative practices of everyday care. Across the four NPT constructs, HCWs described a progression from concerns that disrupted implementation, to priorities that clarified what required strengthening, to pathways that could support more durable integration of TPT into routine services.

### Coherence: From Limited Understanding to Structured Sense-Making

A central concern undermining TPT implementation was the limited awareness and understanding of TPT protocols among HCWs, indicating weak coherence and incomplete understandingat the point of care. Insufficient knowledge and inadequate training impeded effective delivery and reduced providers’ ability to confidently guide patients through treatment, including anticipating and managing side effects. Significantly, these gaps were not merely indicative of individual knowledge deficits; they also highlighted a wider issue in the translation of policy into clear, practical, and standardized guidance for those on the front lines of practice.This mirrors findings from China, where HCW training and public awareness were identified as critical components of TB prevention efforts (21). Our findings extend these reports by showing that even where policy exists, implementation may remain fragile if providers do not encounter TPT as a standardized and coherent part of their daily work.The limited awareness of TPT protocols further reflects a broader weakness in contextual integration(24), where policies are not adequately translated into frontline practice.

In response, HCWs prioritized the need for clearer, standardized, and accessible guidance to support consistent understanding across cadres. The dissemination of SOPs, coupled with improved training and structured communication, was seen as essential to strengthening coherence and enabling shared understanding.

To operationalize these priorities, HCWs proposed actionable pathways including the integration of SOPs into electronic medical record (EMR) systems, the use of automated reminders, and the development of tailored educational materials for providers. Evidence from multi-country studies demonstrates that digital reminders can improve adherence to TB treatment protocols(25), while visual aids and educational tools have been shown to enhance guideline adherence in Ethiopia (26). Regular reinforcement through provider-focused materials and interactive learningplatforms(26,27,29) may further strengthen knowledge dissemination and support sustained coherence in TPT implementation.Collectively, these findings indicate that coherence is enhanced not just through initial training, but also through ongoing, system-supported opportunities to engage with, implement, and reinforce TPT guidance in everyday care.

### Cognitive Participation: Expanding Engagement Beyond Doctor-Centric Models

A key concern affecting cognitive participation was the uneven engagement of HCWs across cadres, with TPT delivery remaining largely doctor-centric. Nurses and pharmacists often improvised ways to engage patients, reflecting commitment in practice but limited recognition in implementation design. This lack of structured engagement limited collective ownership of TPT and weakened implementation sustainability.The concentration of TPT within the responsibilities of doctors led to vulnerabilities, including bottlenecks, discontinuity, and inconsistent patient support throughout various care touchpoints.

HCWs therefore prioritized expanding engagement across the healthcare team by strengthening capacity-building initiatives and fostering shared responsibility in TPT delivery. Continuous, targeted training emerged as a critical need to empower providers with the knowledge and confidence required to actively participate in TPT implementation.The focus on collective accountability is crucial, as the integration of TPT into routine practice relies not just on providers’ comprehension, but also on their perception of its legitimacy and practicality within their professional responsibilities.

The pathways proposed to address these gaps included structured, team-based training approaches and the integration of multimedia resources to enhance both provider and patient engagement. Evidence from Uganda demonstrates that educational videos and printed materials can improve provider knowledge and patient outcomes in TB programs(29). Similarly, incorporating interactive training tools and digital platforms (27,28) may support broader engagement and facilitate sustained participation across cadres. These approaches highlight the importance of moving beyond individual responsibility toward collective, system-wide engagement in TPT delivery.Our findings therefore suggest that successful implementation of TPT necessitates a thoughtful redistribution of ownership among the care team, rather than solely depending on doctor-led initiation.

### Collective Action: Addressing System Constraints and Operational Fragility

Within collective action, chronic understaffing emerged as a critical concern, significantly constraining the operationalization of TPT within routine workflows. Limited workforce capacity disrupted continuity of care, increased patient burden, and created gaps in service delivery, ultimately compromising access to preventive therapy. Similar challenges have been documented in South Africa, where inadequate staffing was identified as a primary barrier to effective TB care (30).In our study, however, staffing constraints were perceived not merely as isolated shortages, but rather as components of a broader pattern of operational fragility. This fragility manifested in a reliance on overstretched personnel, informal workarounds, and inconsistent continuity mechanisms for effective implementation.Beyond operational workload, HCWs repeatedly described medication stock interruptions as a major barrier to implementation. Inconsistent drug availability not only disrupted continuity of care but also weakened provider confidence in recommending TPT. HCWs questioned how preventive therapy could be meaningfully normalized within routine HIV care when medications were intermittently unavailable. These findings suggest that uninterrupted drug supply is not simply a logistical issue, but a prerequisite for provider trust, patient confidence, and sustained implementation(31–33).

To address these challenges, HCWs prioritized strengthening workforce capacity and optimizing service delivery models. Suggested priorities included improving staff allocation, addressing high turnover, and ensuring continuity of care through better workforce planning. HCWs emphasized that institutionalizing TPT processes through standardized documentation systems, routine orientation mechanisms, and formalized workflows could reduce dependence on individual staff members and mitigate disruptions caused by workforce turnover.

Proposed pathways included organizational strategies such as implementing on-call systems, optimizing workforce distribution, and establishing rotating shifts. Additional strategies, such as recruitment and retention initiatives highlighted in the Indian context(34), could be adapted to mitigate workforce instability. Evidence from Bangladesh and the Philippines further underscores how human resource constraints negatively affect TB program performance(35,36), reinforcing the need for comprehensive workforce strengthening strategies.These findings indicate that TPT implementation cannot be stabilized without investments in the workforce infrastructures that enable continuity, consistency, and follow-through.

Beyond staffing, interruptions in TPT supply chains represented another major concern, disrupting both prescribing practices and patient adherence. Similar findings from Kenya highlight the importance of uninterrupted drug supply in maintaining treatment continuity(31). HCWs prioritized strengthening supply chain systems and ensuring consistent availability of medications.This is particularly important for preventive therapies like TPT, as inconsistent access can undermine provider confidence in prescribing and diminish patient trust in the health system’s capacity to maintain care.

To address these issues, pathways included improving supply chain management through digital tracking systems(32,37) and introducing financial support mechanisms such as subsidized treatment (33). These system-level interventions aim to reduce stockouts, enhance program efficiency, and support uninterrupted care delivery.

### Reflexive Monitoring: Adapting Through Experience and Feedback

Reflexive monitoring revealed that HCWs continuously evaluated and adapted TPT implementation in response to patient-level and system-level challenges. A key concern was the impact of supply disruptions and high workload on patient adherence and provider decision-making, often forcing HCWs to prioritize treatment completion over new initiation.

HCWs identified gaps in patient understanding and engagement as critical barriers, with inadequate counselling limiting patients’ ability to adhere to TPT. These findings align with broader evidence emphasizing the importance of patient-provider relationships in sustaining TB treatment adherence (38).

In response, HCWs prioritized strengthening patient-centred counselling and improving communication around TPT benefits and side effects. Enhanced counselling was seen as essential to building patient trust, improving adherence, and mitigating concerns related to pill burden and adverse effects(8,19).Notably, HCWs did not frame counseling as supplementary to implementation; rather, they viewed it as central to whether TPT could be normalized in practice.

Proposed pathways included integrating structured counselling approaches into routine care, supported by dedicated adherence counsellors and improved communication strategies. Evidence from TB programs in Nigeria and Uganda highlights the effectiveness of participatory and stakeholder-engaged approaches in improving implementation outcomes (8,39). These adaptive practices reflected not only commitment and ingenuity among frontline providers, but also the absence of sufficiently embedded institutional supports(11,12,15,38).

### Implications for Policy and Practice

Our findings underscore that TPT implementation is not a static set of barriers but an ongoing process of negotiation, adaptation, and coordination. By systematically incorporating HCWs’ perspectives and contextual realities, programs can identify operational challenges early and develop targeted, context-specific interventions. This participatory approach fosters ownership among frontline providers and enables adaptive recalibration of implementation strategies, ultimately supporting sustainable improvements in TPT delivery.

For effective policy and practice, enhancing the implementation of TPT necessitates efforts that extend beyond merely distributing guidelines. Investment in the establishment of routine practices is essential: this includes clear SOPs, equitable task distribution among various roles, integrated digital reminders, a consistent supply of medications, and monitoring frameworks that ensure TPT is both visible and accountable within standard HIV service delivery. Interventions that address these systemic conditions are more likely to facilitate sustained implementation compared to those that concentrate only on provider knowledge or patient motivation.

Future research should expand this evidence base by exploring patient experiences with TPT through qualitative approaches and evaluating the long-term impact of workforce strengthening and digital health interventions on treatment outcomes. Examining health system contexts in a comparative manner can aid in pinpointing the implementation supports that can be transferred and those that necessitate local adaptation.Addressing these challenges through integrated, context-sensitive strategies will be essential for developing resilient, patient-centered TB prevention systems and advancing progress toward global TB elimination goals.

## CONCLUSION

This study underscores the value of seeking to understand integration or normalization of a new intervention into routine practice. By applying the NPT framework, we show that the successful integration depends on how TPT is understood, shared across cadres, operationalized within workflows, and continuously adapted in response to frontline realities.

Integrating TPT into standard TB/HIV care necessitates comprehensive system-level investments that extend beyond mere training. This includes the establishment of standardized protocols, the implementation of team-based delivery models, the development of reliable supply chains, and the provision of digital or organizational supports that ensure consistent practice. Transitioning from models centered around physicians to a more collaborative approach that includes nurses, pharmacists, and counselors is essential for enhancing implementation and fostering patient engagement.

The ongoing integration relies significantly on incorporating reflexive monitoring within health systems, which encompasses the regular utilization of data, feedback mechanisms, and processes for institutional accountability. Collectively, these strategies can connect policy with practice and facilitate the integration of TPT as a fundamental aspect of HIV care.

## Data Availability

The data underlying the findings of this study cannot be made publicly available because they contain potentially identifiable qualitative information from healthcare providers and public release would compromise participant confidentiality, as restricted by the ethics approvals and informed consent procedures. De-identified data may be made available to qualified researchers upon reasonable request, subject to approval by the relevant Institutional Review Boards and execution of an appropriate data sharing agreement. Requests for access may be directed to the corresponding author.

## List of abbreviations

ART: Antiretroviral therapy
CIDTRAM: CiptoMangunkusumo General Hospital, Indonesia
FGD: Focus Group Discussion
HCW: Healthcare Worker
HIV: Human immunodeficiency virus
IDI: In-Depth Interview
INH: Isoniazid
IPT: Isoniazid Preventive Therapy
MAXQDA: Qualitative Data Analysis Software
NPT: normalization process theory
NTEP: National Tuberculosis Elimination Programme
PLHIV: People living with HIV
RITM: Research Institute for Tropical Medicine, Philippines
RTA: Reflexive thematic analysis
SOP: standard operating procedure
TB: Tuberculosis
TPT: Tuberculosis Preventive Treatment

## Declarations

### Ethics approval and consent to participate

This study received ethical clearance from the Institutional Review Boards of RITM (Approval No. 2022--20), Cipto (Approval No. 22--07--0832) and Johns Hopkins University (Approval No. IRB00335184). Informed consent was obtained from all the participants before they participated in the study. All the study activities were carried out in accordance with the relevant guidelines, including those set forth in the Declaration of Helsinki and the Belmont report.

### Consent for publication

Not applicable

### Availability of data and materials

The datasets generated in this study are not publicly available because of the sensitive and personal nature of the information. Data may be available upon request to the authors, with restrictions following ethical approval.

### Authors’ contributions

PK wrote the initial draft of the manuscript. PK, MDCR, and LS performed data analysis. PK, LS, and CEL contributed to report writing and interpretation of findings. RAHC, MD-L, AGG, RD, MY, AH, EY, and PR critically reviewed the manuscript and contributed to interpretation of the study findings. PK, MDCR, LS, and RAH performed project administration and coordination. MDCR, CEL, RAH, AFP, BJS, MY, AH, EY, AGG, and RD contributed to study implementation, participant recruitment, data collection, and/or data management. EY, AGG, and RD provided site leadership and oversight for study implementation. CJH and JEG provided overall study supervision and critically reviewed and edited the manuscript. All authors read and approved the final manuscript.

## Acknowledgments

We acknowledge the other members of the BEAT TB Asia team from RITM, the Philippines: Dr. Vorognoa Medel Aguilas, Ms. Johanna Beullah, Dr. T. Sornillo, Dr. MarianetteInobaya, and Dr. Aileen Gianan-Gascon; ***JohnsHopkins University***, USA: Dr. Samyra Cox, Dr. Kate Shearer; and ***Johns Hopkins Center forInfectious Disease***, India: Dr. Vidya Mave, Ms. Vedika Potphode. Lastly,we thank all our participants for their time and valuable insights.

## Authors’ information (optional)

Prashant Kulkarni: https://orcid.org/0000-0002-6066-586X

Mark Donald C. Reñosa: http://orcid.org/0000-0002-7414-7174

Bianca Joyce T. Sornillo: https://orcid.org/0009-0001-0023-139X

